# Combination of radiosensitivity index and PD-L1 status predicts overall survival of endometrial cancer patients treated with radiotherapy: A study based on TCGA data

**DOI:** 10.1101/2024.10.27.24316237

**Authors:** Shaohua Xu

## Abstract

Identifying biomarker for the clinical outcome of patients with endometrial cancer (EC) after radiotherapy is helpful to optimize treatment. The predictive efficacy of the combination of radiosensitivity index (RSI) and programmed cell death ligand 1 (PD-L1) should be investigated. The endometrial cancer cohort contained 225 patients received radiotherapy, and 285 patients who did not receive radiotherapy was downloaded from The Caner Genome Atlas (TCGA) and divided into radiosensitive (RS) and radioresistant (RR) groups according RSI. The EC patients were further classified into PD-L1-high and PD-L1-low groups according to the median value of CD274 mRNA expression. Cox proportional hazards regression and Kaplan-Meier analysis for overall survival were performed. The differently expressed analysis, gene set enrichment analysis and immune cell infiltration were performed. Among EC patients in the TCGA dataset, a group characterized by radioresistance and low PD-L1 status (PD-L1-high-RR group) was identified. Kaplan-Meier analysis showed EC patients in the RR-PD-L1-low group had significantly worse overall survival than those in the other groups when received radiotherapy (p=0.0006). Cox proportional hazards regression showed patients in the RR-PD-L1-low group were significantly associated with a poor of overall survival (HR=3.79; 95% CI=1.43-6.23; p=0.007) when received radiotherapy. MAPK signaling pathway was differently enriched in RR-PD-L1-high group. CD8^+^ T cell and M1 macrophage were lower infiltration in RR-PD-L1-high group, whereas M2 macrophage was more infiltration in RR-PD-L1-high group. The combination of RSI and PD-L1 may serve a predictive tool to identify EC patients who benefit from radiotherapy, especially combined immunotherapy.

## 1. Introduction

Endometrial cancer (EC) is a group of epithelial malignant tumors occurring in the endometrium, and ranks as the third most frequently diagnosed cancer and the fourth leading cause of cancer death in women worldwide [1]. There have an estimated 69,950 new cases and 12,550 deaths in the United States in 2022 [2]. Approximately 67% of patients with EC are found at an early stage, and an overall 5-year survival rate can reach 80% [3]. However, the recurrence rate of patients with advanced endometrial cancer (stage III and stage IV) is high, and its prognosis is poor and the overall 5-year survival rate is only 20%-40% [3].

Radiotherapy (RT) is an important treatment for endometrial carcinoma. Prior studies reported that early stage EC patients received adjuvant radiation therapy had improved overall survival in higher-risk subgroups and a local control advantage in the absence of a survival benefit for adjuvant radiation over observation [4,5]. Although radiotherapy can reduce the local recurrence rate of endometrial cancer, it do not make a large contribute to improve the 5-year survival rate and tumor-free survival time of patients [4].

Tumor immunotherapy is a treatment method to control and remove tumors by inducing and augmenting anti-tumor immune response to the body [6]. In gynecological malignancies, the U.S.Food and Drug Administration (FDA) has approved immune checkpoint inhibitors ( ICIs ) monotherapy or in combination with other targeted drugs for the treatment of metastatic or recurrent endometrial cancer with specific biomarkers [7]. Although immunotherapy can produce a lasting response, the response rate of ICIs monotherapy to solid tumors is usually low, and many patients eventually develop secondary resistance [8]. The combination of immunotherapy and radiotherapy is being tested clinically to target multiple defects in the immune cycle and intrinsic changes in cancer, thereby improving anti-cancer efficacy. Some studies reported that immunotherapy, especially immune checkpoint blockade, may reduce tumor hypoxia via modulating the tumor microenvironment [9]. These findings may suggest the potential opportunity for immune checkpoint blockade therapy to increase radiosensitivity. Multiple clinical trials showed an increased clinical significance and survival advantage of patients with various cancers in the combination of immune checkpoint inhibitors and radiotherapy, including non-small cell lung cancer[10], hepatic cancer[11] and rectal cancer[12]. Currently, several clinical trials of combined immunotherapy and radiation for metastatic or recurrent gynecologic cancer are ongoing [13].

One of the important factors of immunotherapy and radiotherapy regimens is to screen more proper and accurate biomarkers for accurately finding the best beneficiaries. It makes sense that not all patients would have the same benefit for RT in combination with immunotherapy. Currently, there has no robust biomarker that can aid to assess which EC patients may need treatment with combination therapy and, which EC patients are not benefiting from combination therapy. Thus, the discovery of predictive and evaluative biomarkers is an important requirement to develop clinical treatment strategies for EC patients to a combination of radiotherapy with immune checkpoint blockade therapy. Previously, a radiosensitivity index (RSI) containing 10 genes was developed to predict cancer patients which are sensitivity to radiotherapy [14]. The RSI has been validated in several cancer types to predict clinical outcome of patients treated with radiotherapy, including head and neck cancer[15], breast cancer[16], pancreatic cancer[17], colon cancer[18] and glioblastoma[19]. In 2019, ASTRO reported a study used RSI to predict the risk of pelvic failure after adjuvant radiotherapy in endometrial cancer, showing that patients with radiation resistance (RR) were more likely to have pelvic failure and worse pelvic failure-free survival than patients with radiation sensitivity (RS)[20]. Furthermore, for immunotherapy, programmed cell death ligand 1 (PD-L1) testing is the most recognized indicator of therapeutic response[21].

In the present study, we hypothesize that integrating radiosensitivity index gene signature and PD-L1 expression is able to predict the clinical outcome of EC patients treated with radiotherapy, particularly combined immunotherapy. The combination of RSI and PD-L1 can be used as a predictive tool to identify EC patients who can benefit from radiotherapy, especially combined immunotherapy.

## 2. Methods

### 2.1 TCGA data

The gene expression RNAseq level 3 data of GDC TCGA Endometrioid Cancer (UCEC) was downloaded from UCSC Xena[22]. The gene level expression was measured in Fragments per Kilobase of transcript per Million mapped reads (FPKM) according GDC mRNA quantification analysis pipeline (https://docs.gdc.cancer.gov/Data/Bioinformatics_Pipelines/Expression_mRNA_Pipeline/). A total of 583 samples were available in the dataset (07-21-2019 version). Then, clinical information and survival data were also achieved from UCSC Xena. After matching the clinical information and survival data and removing samples of metastatic tumors, 225 patients who underwent radiotherapy and 285 patients who didn’t undergo radiotherapy were included and further analyzed.

### 2.2 Radiosensitivity index and PD-L1 status

Based on previously published RSI linear regression algorithm[14]: RSI score = 0.0128283 * cJun - 0.0098009 * AR + 0.0254552 * STAT1 - 0.0017589 * PKC - 0.0038171 * RelA + 0.1070213 * cABL - 0.0002509 * SUMO1 - 0.0092431 * PAK2 - 0.0204469 * HDAC - 0.0441683 * IRF1, we calculated the RSI score of each patient. Then, all patients was divided into two groups: radiosensitive (RS) group and radioresistant (RR) group according to cut-off which acquired using the 75th percentile for RSI score[23]. The cut-offs were 2.948 and 3.329 for RT patients and no RT patients, respectively. The patients whose RSI score were higher than cut-off were included in RR group, whereas the patients whose RSI score were lower than cut-off were included in RS group. On the other hand, CD274 gene expression was considered as PD-L1 surrogate. According to median FPKM value of the CD274 gene, patients were divided into two groups: PD-L1-high and PD-L1-low groups. The patients whose CD274 were higher than median FPKM value of the CD274 in PD-L1-high group, whereas the patients whose CD274 were lower than median FPKM value of the CD274 included in PD-L1-low group. Finally, four subgroups: RR-PD-L1-high, RR-PDL-1-low, RS-PD-L1-high and RS-PD-L1-low groups were defined according to RSI score and PD-L1 status.

### 2.3 Bioinformatics analysis

The differentially expressed genes (DEGs) between the RR-PD-L1-low group and other group were identified using DESeq2 R package[24]. A cutoff point to identify the significant DEGs was that p value is less than 0.05 and absolute log fold change more than 1. Gene set enrichment analysis (GSEA) with Kyoto Encyclopedia of Genes and Genomes (KEGG) pathway was performed on the DGEs using clusterProfiler R package[25]. Immune cells infiltrated in each group were estimated on TIMER2.0 web server [26] (http://timer.cistrome.org/) using CIBERSORT [27], xCELL[28], TIMER[29], and EPIC[30] algorithms.

### 2.4 Statistical analysis

The difference of clinical characteristics between RT and non-RT group, RR and RS group was test using chi-square analysis. The CD274 mRNA expression level was compared between RR and RS group using wilcoxon signed-rank test. Univariate and multivariate logistic regression analysis was carried out to evaluate which clinical factors were related to PD-L1 status. Univariate and multivariable Cox proportional hazards regression was performed to evaluate association with overall survival. Kaplan-Meier survival curve for overall survival with log-rank test was performed. Continuous variables were compared with wilcoxon signed-rank test or t test, while categorical variables were compared with chi-square test. A p < 0.05 was considered statistically significant. All statistical analysis were performed by R version 4.0.1.

## 3. Results

### 3.1 RSI for predicting overall survival of EC patients treated with radiotherapy

A total of 510 patients (225 patients who receive radiotherapy and 285 patients who didn’t receive radiotherapy) were brought into this study from TCGA database. Patients’ characteristics are summarized in Table 1. To validate the RSI applied to the TCGA UCEC datasets, the patients were divided into radiosensitive (RS) group and radioresistant (RR) group according to the cut-off of RSI score and the Kaplan-Meier curves for overall survival (OS) is displayed in Fig.1. RSI statistically significantly stratified the RT-treated patients for OS (log-rank test, p=0.004). Meanwhile, the differences in OS were not observed between RR group and RS group who did not receive radiotherapy (log-rank test, p=0.082).

**FIGURE. 1.**
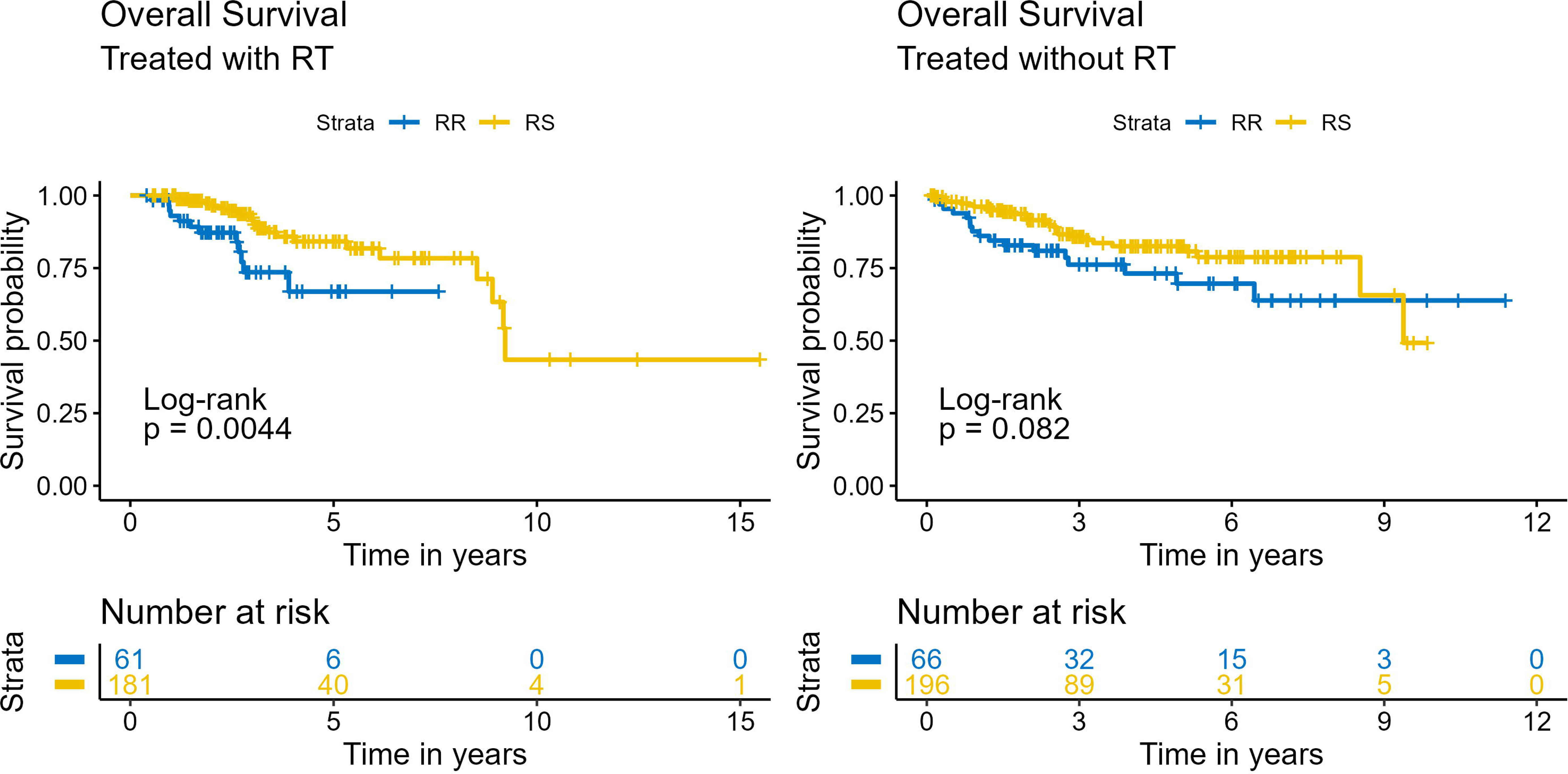
Survival analysis according to radiosensitivity index. Kaplan-Meier survival curves of overall survival from endometrial cancer patients treated with or without radiotherapy.

**Table 1.**
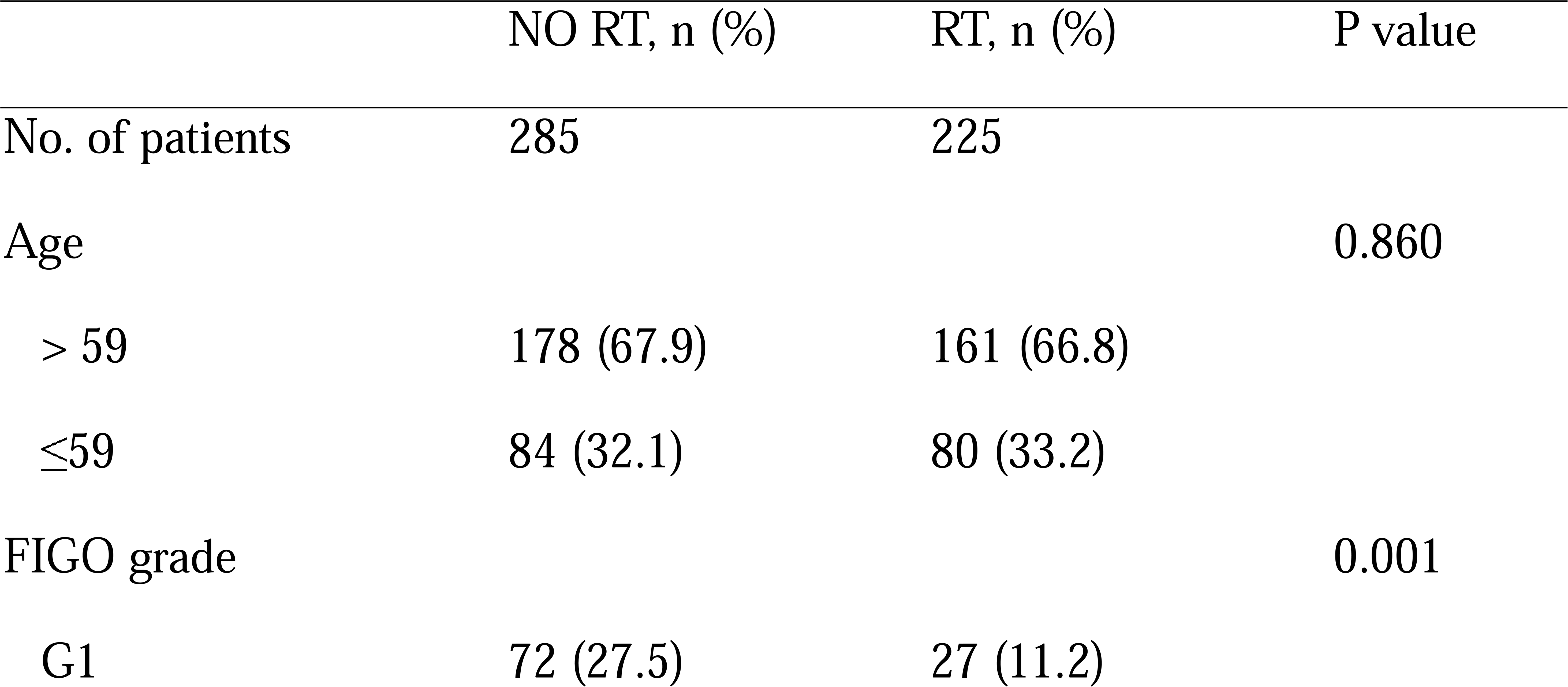
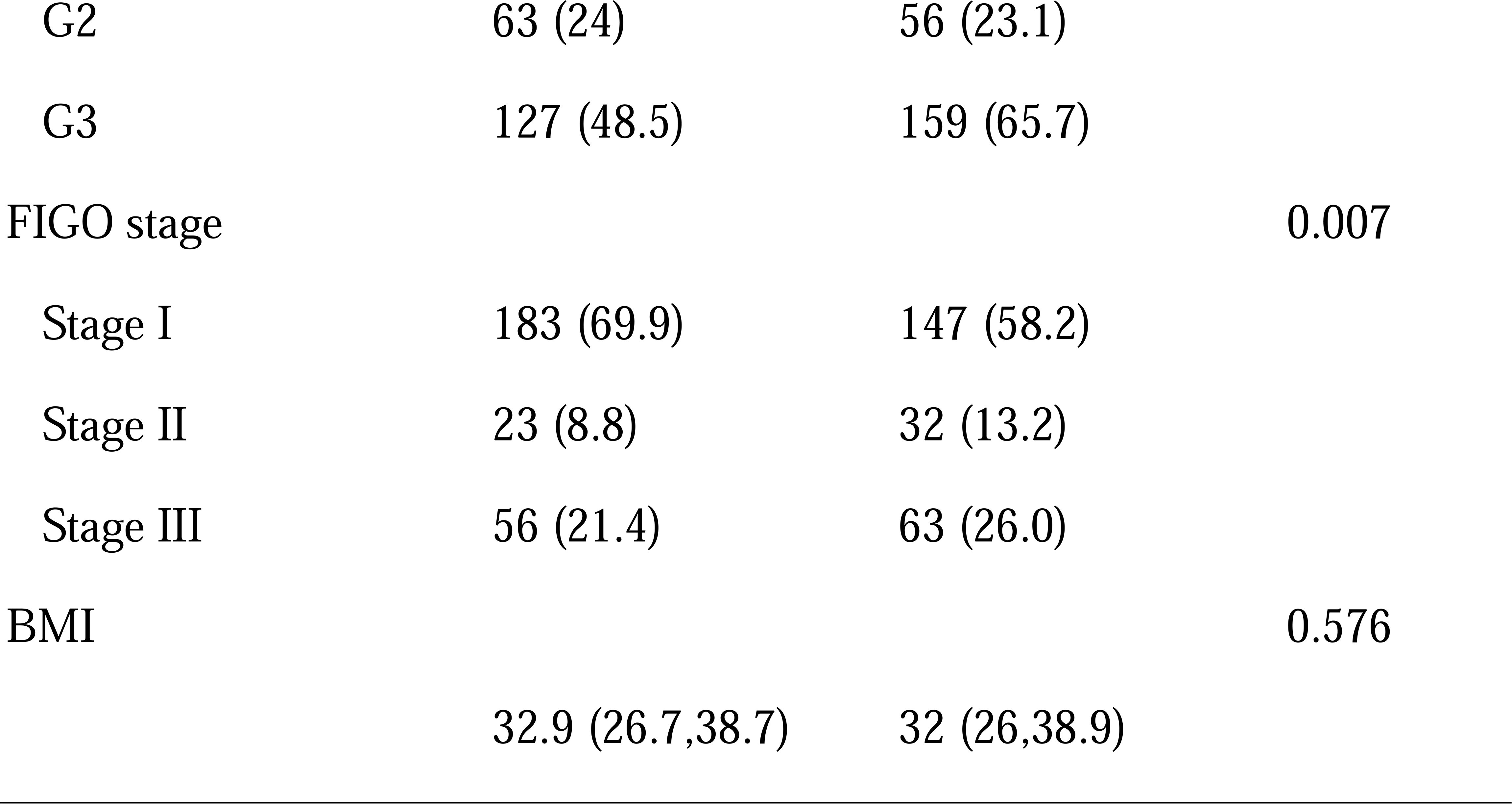
Clinical characteristics of the overall patients.

### 3.2 PD-L1 status

Based on the median value of the CD274 gene expression, RT-treated patients and patients who didn’t receive RT were divided into two groups: PD-L1-high group and PD-L1-low group, respectively. We compared the PD-L1 status of patients treated with RT between in the RR and RS group. There was significantly different distribution of PD-L1 status between the RR and RS group. The CD274 mRNA expression level in the RS group was significantly lower in the RR group (p=0.026) (Fig. 2). Above results showed RSI score was associated with PD-L1 status.

**FIGURE. 2.**
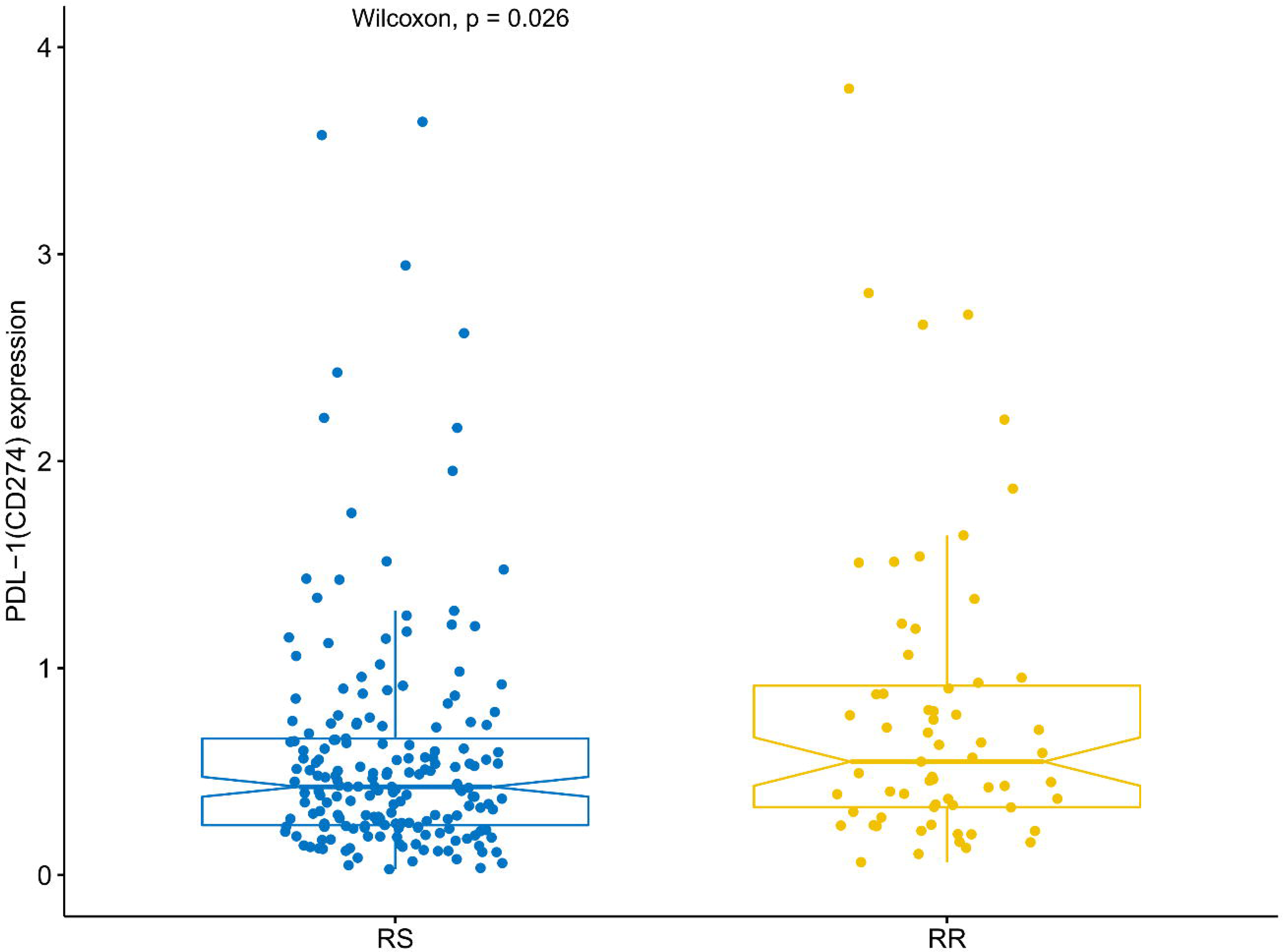
CD274 mRNA expression level was compared between radiosensitivity (RS) group and radioresistant (RR) group.

### 3.3 Survival analysis according to RSI and PD-L1 status

To evaluate whether the RSI score and PD-L1 status were associated with overall survival for patients received RT, univariate and multivariate Cox proportional hazard regression analysis were performed for RT-treated patients (Fig. 3A). Univariate analysis indicated that patients in the RS group significantly correlated with an improved OS (HR=0.35; 95%CI=0.17-0.75; p=0.006), and PD-L1 status was not associated with OS (p=0.078). Multivariate analysis showed RSI was the strongest factor and patients received RT in RS group was significantly associated with improved OS (HR=0.31; 95%CI = 0.12-0.77; p=0.011), taking into account age, FIGO grade, FIGO stage, and BMI.

**FIGURE. 3.**
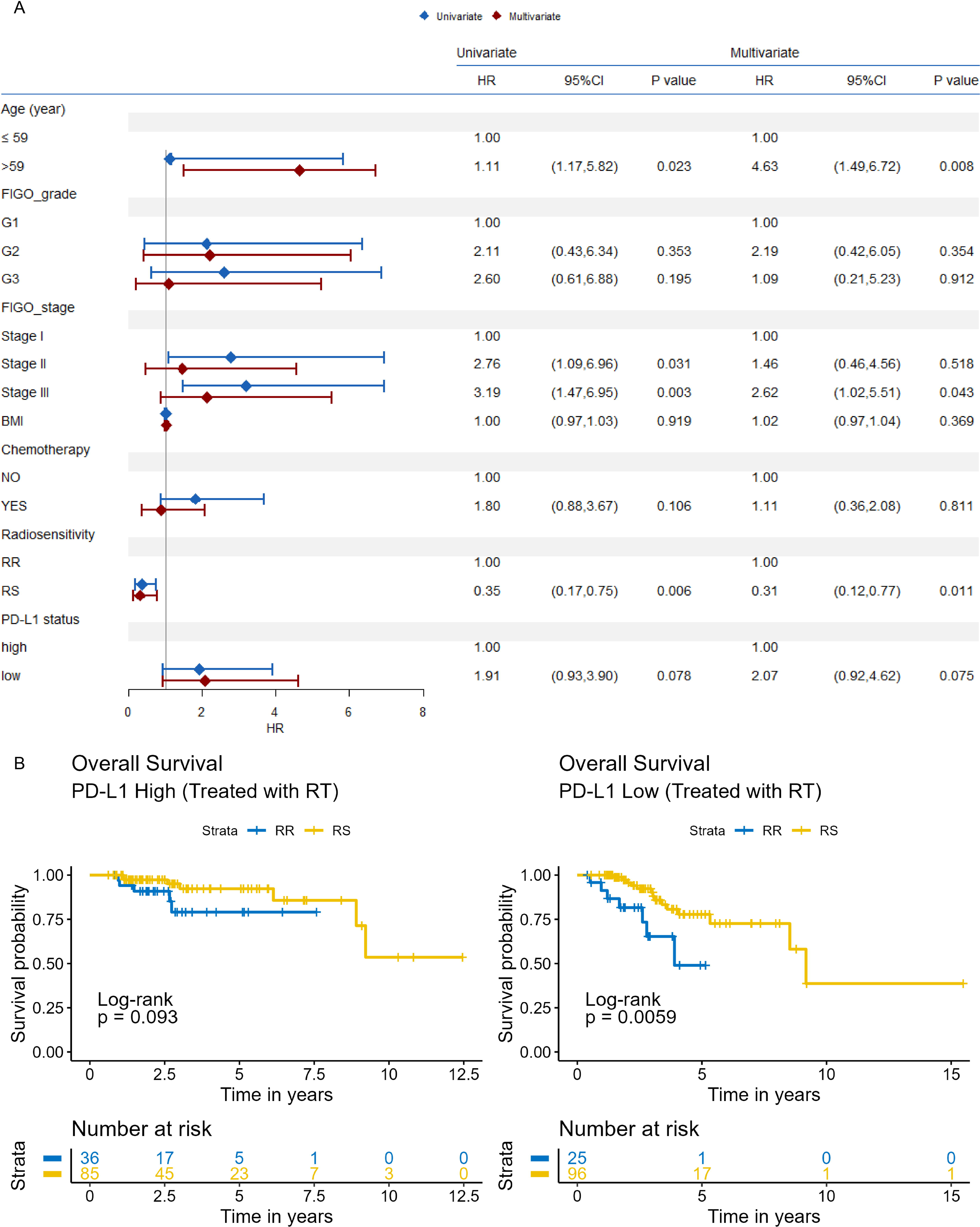
Survival analysis according to radiosensitivity index and PD-L1 status. A. Univariate and multivariate Cox proportional hazard regression analysis were performed for RT-treated patients. B. Kaplan-Meier survival curves of overall survival were plotted for endometrial cancer patients treated with radiotherapy according to radiosensitivity index and stratified to PD-L1-high and PD-L1-low group.

The Kaplan-Meier curves displayed that on stratification by PD-L1 status, RS patients who received radiotherapy had better OS than RR patients in the PD-L1-low status (log-rank test, p = 0.0059). By contrast, there was not differences in OS between patients in the RR and RS group who received radiotherapy in the PD-L1-high status (log-rank test, p=0.093). The results were shown in Fig. 3.B.

### 3.4 Combination of RSI and PD-L1 status for predicting clinical outcome of EC patients treated with radiotherapy

In order to further determine whether the RSI score and PD-L1 status is association, we conducted a subgroup analysis in the whole patients. Based on RSI score and PD-L1 levels, the whole patients were divided into four groups: RR-PD-L1-high, RR-PDL-1-low, RS-PD-L1-high and RS-PD-L1-low group. Kaplan-Meier curves were subsequently plotted for the four subgroups (Fig. 4). RT-treated patients in the RR-PD-L1-low group had the significantly shortest OS than those in the other three subgroups (log-rank test, p=0.002). Meanwhile, patients not received RT in the four subgroup had no significant difference for OS (log-rank test, p=0.38). Then, RR-PD-L1-high, RS-PD-L1-high and RS-PD-L1-low group were merged into one group: other group. Kaplan-Meier curves displayed that OS of patients treated with RT in other group was superior compared with subjects in RR-PD-L1-low group (log-rank test, p=0.0006), while OS was not significantly different between with this two groups when RT was not given (log-rank test, p=0.22) (Fig. 4).

**FIGURE. 4.**
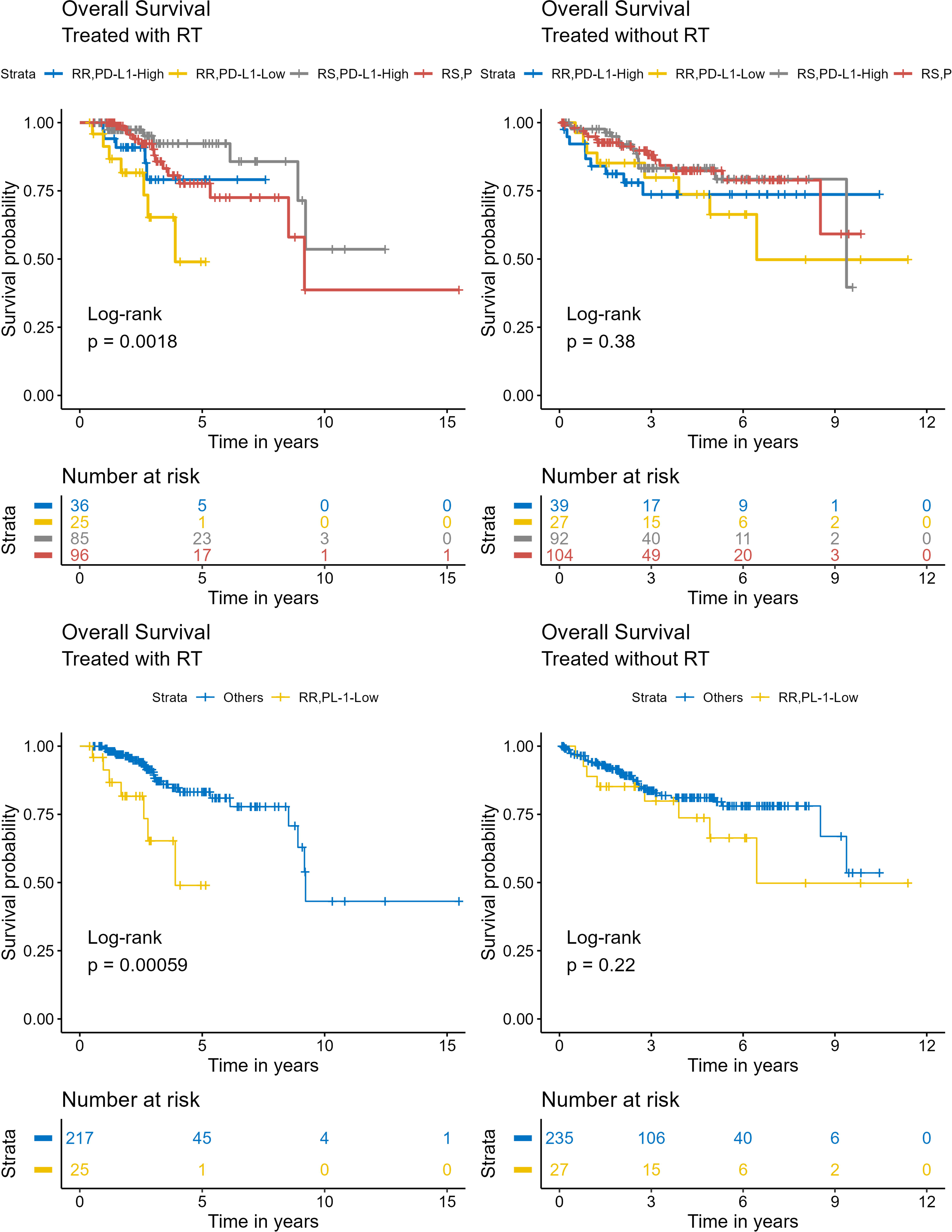
Kaplan-Meier survival curves of overall survival were plotted endometrial cancer patients treated with or without radiotherapy according to combining radiosensitivity index and PD-L1 levels.

To further explore the prognostic value of RSI score and PD-L1 status when combined with clinical characteristics as age, FIGO grade, FIGO stage and BMI on overall survival of RT-treated patients, multivariate Cox proportional hazard regression analysis was carried out. The results showed that RSI combined PD-L1 status was able to stratify RT-treated patients for OS independent of patient age, FIGO grade, FIGO stage, and BMI, and the patients in RR-PD-L1-low group was significantly associated with worse OS than the other group (HR=3.79; 95% CI=1.43-6.23; p=0.007) (Fig. 5).

**FIGURE. 5.**
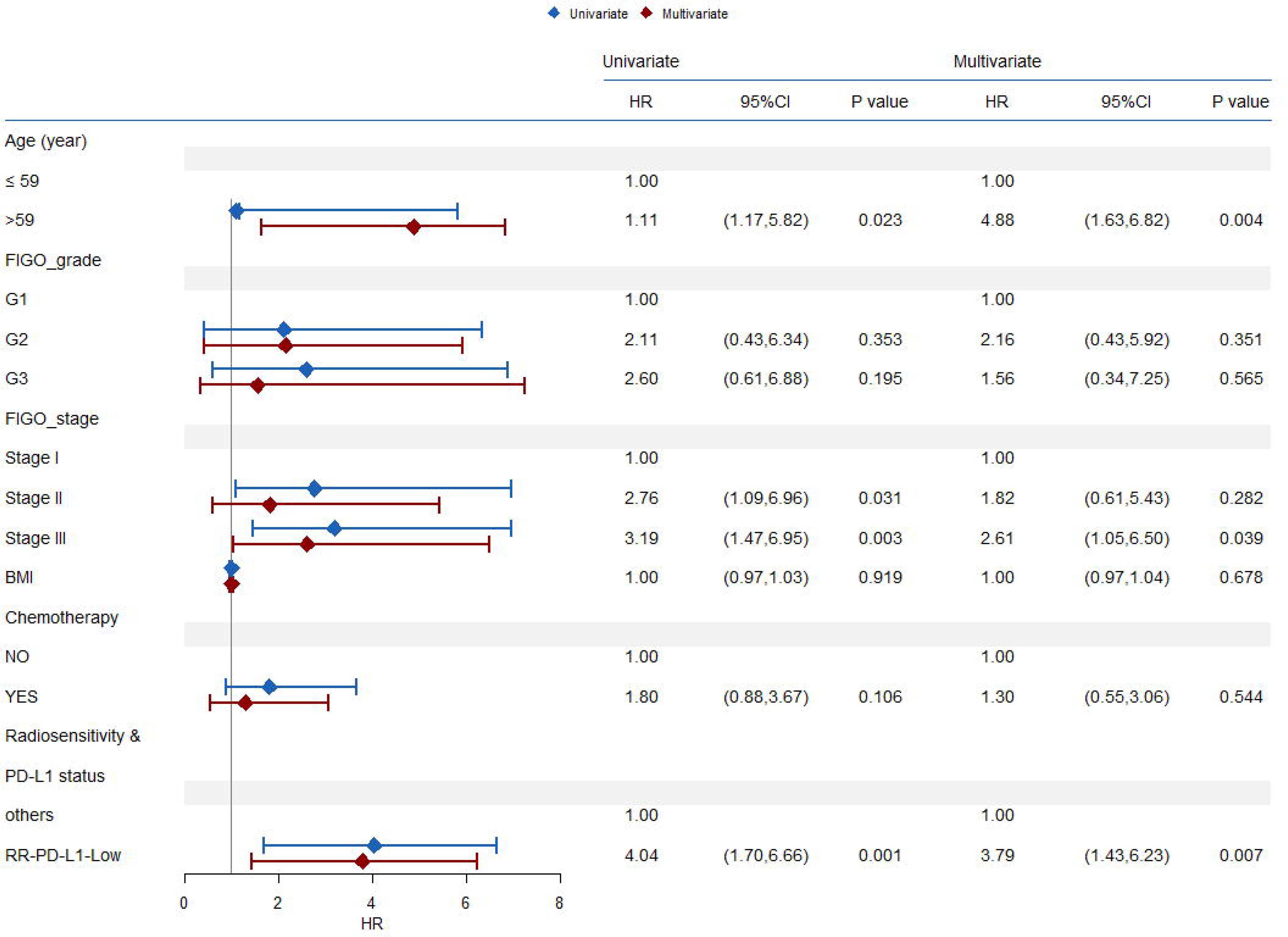
Cox proportional hazard regression analysis were performed for RT-treated patients in PD-L1-low-RR group and other group.

### 3.5 Differentially expressed genes identification between RR-PD-L1-high group and other group

A total of 2089 differentially expressed genes were screened between other group and RR-PD-L1-low group, among which 1728 genes were down-regulation and 361 genes were up-regulation (Fig. 6A). To explore the functions of differentially expressed genes, the KEGG pathway gene set enrichment analysis (GSEA) was then performed. MAPK signaling pathway, Wnt signaling pathway, Chemical carcinogenesis-receptor activation, and Ras signaling pathway were differentially enriched in RR-PD-L1-high group (Fig. 6B-C).

**FIGURE. 6.**
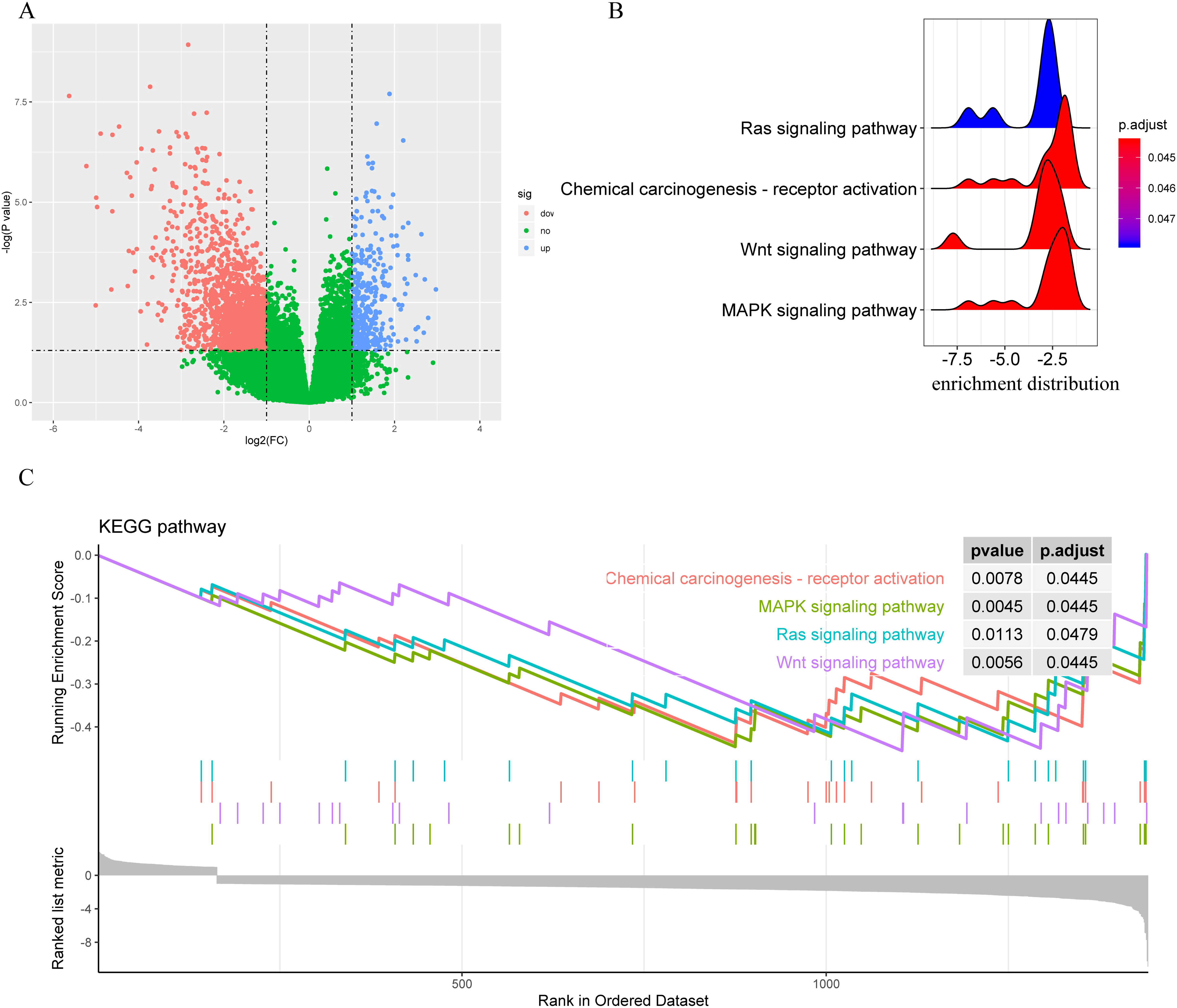
identification of differentially expressed genes and Gene Set Enrichment Analysis. A. The volcano plot for differentially expressed genes. B. The gene set enrichment analysis for differentially expressed genes. C. Enrichment plots from gene set enrichment analysis.

### 3.6 Estimation of immune cell infiltration

To identify the specific immune cell types infiltrated into RR-PD-L1-low group and other group and according to RSI and PD-L1 status, immune cell infiltration was estimated using four methods. The immune cell infiltration between RR-PD-L1-low group and other group was shown in Fig. 7. The CIBERSORT, XCELL, TIMER, and EPIC analysis showed CD8^+^ T cell was lower infiltration in tumor samples of the RR-PD-L1-low group than those in other group. CIBERSORT and xCELL analysis showed the RR-PD-L1-low group had less infiltration of M2 macrophage than other group. The RR-PD-L1-low group had more naive B cell infiltration and less monocyte infiltration in comparison of other group through CIBERSORT method.

**FIGURE. 7.**
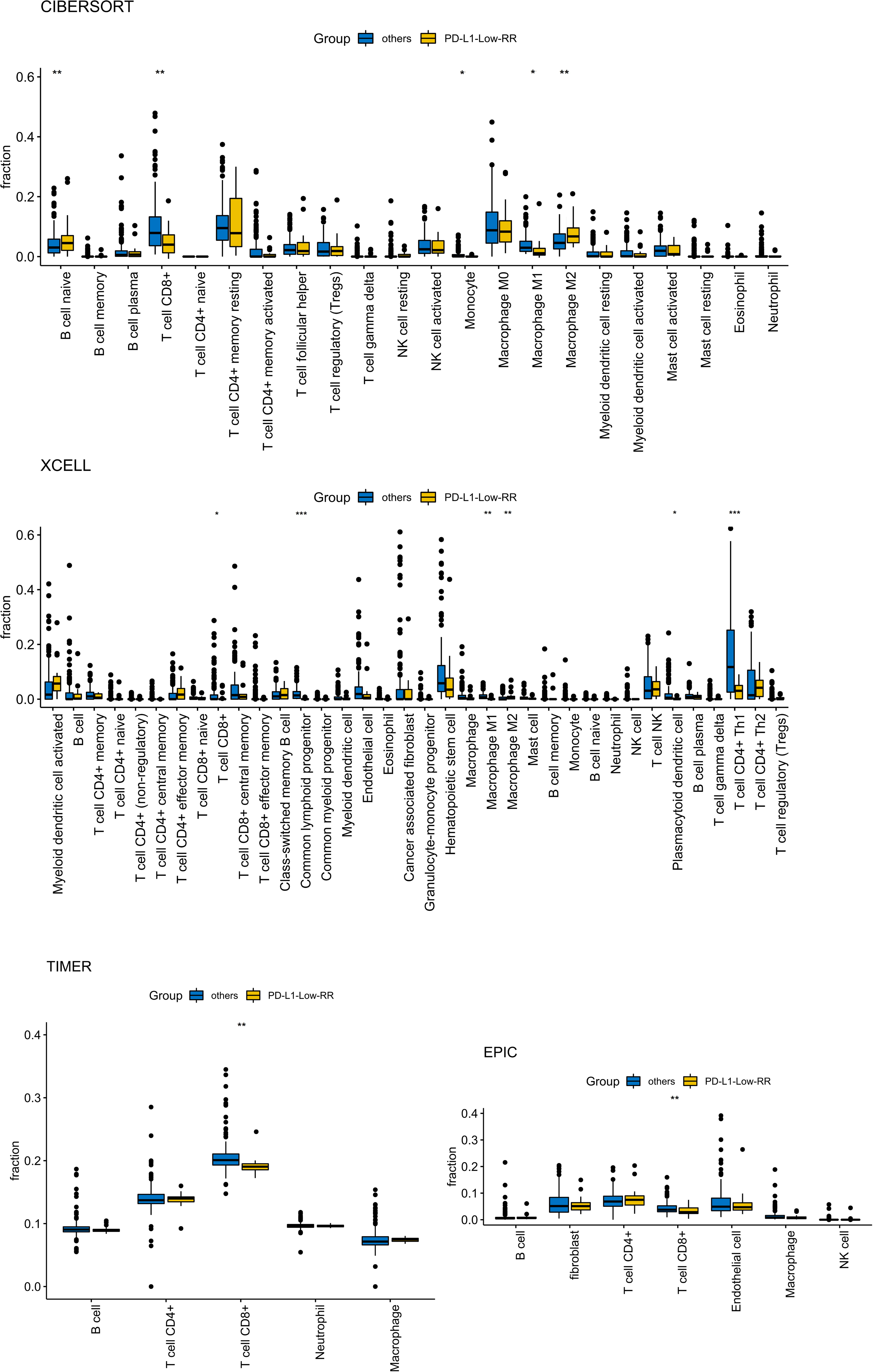
The distribution of immune cell infiltration level between PD-L1-low-RR group and other group.

## 4. Discussion

Recently, several clinical trials of radiotherapy combined with immunotherapy, especially immune checkpoint inhibitors therapy for various cancers showed an increased clinical significance and survival advantage of patients [10–12]. One of the important factors for selecting immunotherapy and radiotherapy regimens is to discover more appropriate and robust biomarkers to accurately find the best beneficiaries. Biomarkers such as PD-L1 have certain guidance for immunotherapy, but there are also some limits. Considering multiple factors and exploring more biomarkers may help the clinic to better select the patients who benefit from combined treatment. In this study, we proposed that integrating radiosensitivity index gene signature and PD-L1 expression could predict clinical outcome of EC patients treated with radiotherapy, particularly when combined with immunotherapy.

We first validated radiosensitivity index gene signature in an EC cohort from TCGA database to determine that the RSI could predict prognosis of EC patients treated with RT. We observed that on stratification by RSI score, RS patients had better OS than RR patients when patient received radiotherapy. Meanwhile, the RT-patients in the RS group significantly correlated with an improved OS compared to RR group was also seen in results of Cox proportional hazards regression. These results suggested the RSI could predict clinical outcome of EC patients who are treated with radiotherapy.

At present, clinical trials of radiotherapy combined with immune checkpoint blockade are underway in patients with metastatic or recurrent gynecologic cancer. Understanding the immunological characteristics of EC may be a key step before designing clinical studies.Therefore, we then investigated the distribution of PD-L1 gene expression, relationship between RSI score and PD-L1 status, and association of RSI, PD-L1 status and OS in EC patients. We found that PD-L1 expression level in the RS group was significantly lower in the RR group and RS group was associated with PD-L1-low group. Published research showed that high grade EC patients expressed higher PD-L1 level than low grade EC patients [31]. Therefore, our findings were consistent with the previous study. Then, the Kaplan-Meier analysis showed that RS patients who received radiotherapy had better OS than RR patients in the PD-L1-low group whereas RR patients treated with RT was not different from RS patients for OS in the PD-L1-high group. Furthermore, the EC patients were divided into four groups: RR-PD-L1-high, RR-PDL-1-low, RS-PD-L1-high and RS-PD-L1-low group according to a combination of RSI score and PD-L1 expression levels. Patients in the RR-PD-L1-low group had the significantly shortest OS than those in the other three subgroups when treated with RT, whereas patients not received RT in the four subgroup had not significant difference for OS. Moreover, Cox regression analysis also showed RR-PD-L1-low group was significantly associated with a worse OS. These results indicated the a combination of RSI gene signature and PD-L1 expression level should predict clinical outcome of EC patients treated with radiotherapy, and can be as a powerful biomarker for identifying specific EC patients who benefit from radiotherapy, particularly combined with immunotherapy.

To identify genomic feature and immunological feature of tumor samples in the RR-PD-L1-low group, we performed KEGG pathway GSEA and estimation of immune cell infiltration. We found MAPK signaling pathway, Wnt signaling pathway and Ras signaling pathway were differentially enriched in RR-PD-L1-high group. Previous studies showed MAPK signaling pathway, Wnt signaling pathway and Ras signaling pathway were associated with radioresistance to cancer radiotherapy, and inhibition of these pathways could enhance sensitivity to radiotherapy [32–34]. Moreover, we found that CD8^+^ T cell, M1 macrophage and monocyte were lower infiltration in tumor samples of the RR-PD-L1-high group than those in other group, whereas M2 macrophage was more infiltration in tumor samples of the RR-PD-L1-high group than those in other group. This results indicated the tumor samples of patients in RR-PD-L1-high group were immune exhaustion and immunosuppression status. In radiation therapy, CD8^+^ T cells were grouped in local tumor microenvironment resulting a cytotoxic immune response against tumor cells, and patients with more CD8^+^ T cells aggregated in local microenvironment are more sensitivity to radiation [35,36]. And, utilizing CD8^+^ T cell response at tumor microenvironment plays a critical role in caner immunotherapy[37]. M2 macrophages could induce radioresistant to head and neck cancer through secreting human heparin-binding epidermal growth factor[38]. Whereas, M1 macrophages infiltration in the tumor microenvironment increased in sensitivity to radiotherapy[39]. So, patients in the RR-PD-L1-high group had worse clinical outcome when treated with radiotherapy may be partly interpreted by the genomic feature and immune infiltration feature of tumor samples.

## 6. Conclusions

In this study, we sought to test the RSI and PD-L1 status as a predictive biomarker for prognosis in the EC cohort of the TCGA dataset. We analyze relationship among radiosensitivity index and PD-L1expression in EC patients that may contribute to identify specific EC patients who benefit from radiotherapy, particularly combined with immune checkpoint blockade therapy. Based on combination of RSI and PD-L1 level, we discovered a specific patient group (RR-PD-L1-low group) that might be not benefit from a combination of radiotherapy and immunotherapy.

## Data Availability

All data produced in the present study are available upon reasonable request to the authors

## ACKNOWLEDGMENTS

We sincerely thank the TCGA Research Network (http://cancergenome.nih.gov/) for providing patient datasets.

## COMPLIANCE WITH ETHICAL STANDARDS

Not applicable

## DATA AVAILABILITY STATEMENT

The datasets used and/or analysed during the current study are available from the corresponding author on request.

